# Impact of Culture-Bases Family Empowerment on Anemia Prevention Behavior in Pregnant Women: A Systematic Review

**DOI:** 10.1101/2025.09.10.25335498

**Authors:** Jelita Siska Herlina Hinonaung, Esti Yunitasari, Moses Glorino Rumambo Pandin

**Affiliations:** Doctoral Program of Nursing, Faculty of Nursing, Universitas Airlangga, Jl. Dr. Ir. H. Soekarno, Mulyorejo, Kec. Mulyorejo, Surabaya, East Java 60115; Departement of Health, Politeknik Negeri Nusa Utara, Jl. Kesehatan No. 1, Kec. Tahuna, Sangihe Islands Regency, North Sulawesi 95811; Faculty of Humanities, Universitas Airlangga, Jl. Dr. Ir. H. Soekarno, Mulyorejo, Kec. Mulyorejo, Surabaya, East Java 60115

**Keywords:** family support, cultural, nursing, iron deficiency, pregnancy, prevention

## Abstract

**Background:** Anemia is a major public health problem worldwide, especially among pregnant women. This is because the overall iron requirements during pregnancy are significantly higher than those in non-pregnant women. This systematic review aimed to assess the impact of culture-based family empowerment on the prevention of anemia in pregnant women. This systematic review was conducted according to the PRISMA guidelines. The search was conducted in Scopus, PubMed, Web of Science (WoS), Science Direct, and EBSCO databases from 2020 to 2025. The included articles were in English, full-text, and not review articles. Of the total 1419 records screened, the last six studies were included in the systematic review. The results showed that culture-based family empowerment increased hemoglobin levels, family support, and behavior. Conclusion: The family empowerment model, when combined with cultural principles, significantly impacts anemia prevention behavior in pregnant women. This approach not only increases hemoglobin levels but also increases family support and encourages positive behavioral changes. Health workers can adopt this model to optimize anemia prevention strategies and improve maternal health outcomes.

## INTRODUCTION

Anemia is a major public health problem worldwide, particularly in developing countries.(Rahman et al., 2022; World Health Organization, 2025) This is because the overall iron requirement during pregnancy is much higher than in non-pregnant women. Globally, 37% (32 million) of pregnant women experience anemia.(World Health Organization, 2025) The Indonesian Health Survey (SKI) reported that 27.7% of pregnant women experience anemia.(Kementerian Kesehatan Republik Indonesia, 2023) Anemia in pregnancy remains a leading cause of pregnancy complications, premature birth, low birth weight, and maternal and infant mortality.(Lewkowitz & Tuuli, 2023; Stephen et al., 2018)

Efforts to prevent anemia have been implemented, such as administering iron tablets, increasing nutritional intake, and providing health education.(Abdulsalam et al., 2025; Gibore et al., 2021; Lewkowitz & Tuuli, 2023; Rahman et al., 2022; Wambes, 2025) However, the success of these programs is often hampered by low compliance among pregnant women, lack of family support, and differences in cultural values that influence health behaviors.(Bhanbhro et al., 2020; Ngotie et al., 2022; Triharini et al., 2021)

The family is the primary means of socialization for individuals to learn about and understand their culture and values. As the smallest unit in society, families play a crucial role in maternal health decision-making. Families can function as agents of change, socializing the inheritance of cultural values, which can be acquired through learning.(Graves & Shelton, 2007)

The Transcultural Nursing approach developed by Madeleine Leininger emphasizes the importance of considering cultural values, beliefs, and practices in providing nursing care.(Leininger & McFarland, 2002; McFarland et al., 2012) Thus, interventions aligned with the family’s cultural background can increase the acceptance and sustainability of healthy behaviors in pregnant women. Therefore, this systematic review aimed to assess the impact of culture-based family empowerment on anemia prevention behaviors in pregnant women.

## METHODS

### Study Design

The process of literature screening, including, and reporting was based on the guidelines of PRISMA.(Page et al., 2021) The search was conducted in Scopus, PubMed, Web of Science (WoS), Science Direct, and EBSCO databases from 2020 to 2025.

### Inclusion and Exclusion Criteria

The impact of culture-based family empowerment was systematically observed in pregnant women’s anemia prevention behaviors. The inclusion criteria were as follows: research study, study conducted in a group of pregnant women, and full text of the study published in English. The exclusion criteria were as follows: studies conducted in animal models and article reviews.

### Searching Strategy

The literature search used Scopus, PubMed, Web of Science (WoS), Science Direct, and Ebsco databases from 2020 to 2025 using English. The keywords in the search were (pregnant women OR pregnancy) AND (family empowerment OR family support) AND (cultural) AND (prevention OR intervention OR management OR health promotion) AND (anemia OR iron deficiency OR hemoglobin).

The identification, screening, and inclusion procedures of the studies available in Scopus, PubMed, Web of Science (WoS), Science Direct, and EBSCO databases are presented in Figure 1. The entire process was conducted by two independent researchers, initially based on the title and abstract, followed by the procedure based on the full text of the study. In case of any disagreement between the assessing researchers, the third researcher was consulted to resolve the issue. Full texts of the studies were obtained from electronic databases or university libraries.

**Figure 1.**
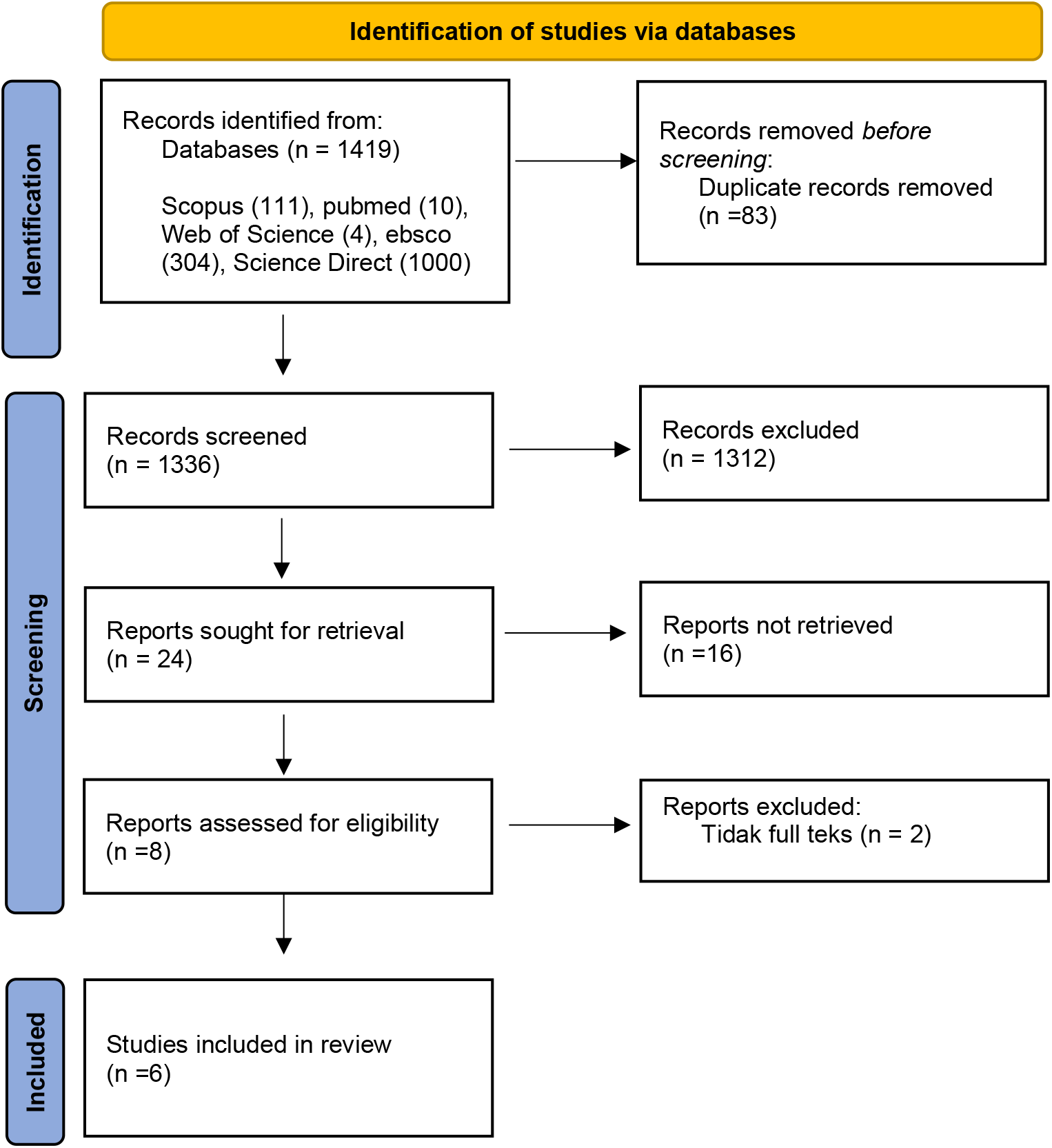
The identification, screening, and inclusion procedures of the studies were available in Scopus, PubMed, Web of Science (WoS), Science Direct, and EBSCO databases.

### Procedure of Data Extraction

Data extraction was conducted by two independent researchers. In case of any disagreement between the assessing researchers, the third researcher was consulted to resolve the issue. All necessary information was obtained from the full texts of the studies.

## RESULT

The results of the research articles on the impact of culture-based family empowerment on anemia prevention behavior in pregnant women are presented in Table 1.

**Table 1.**
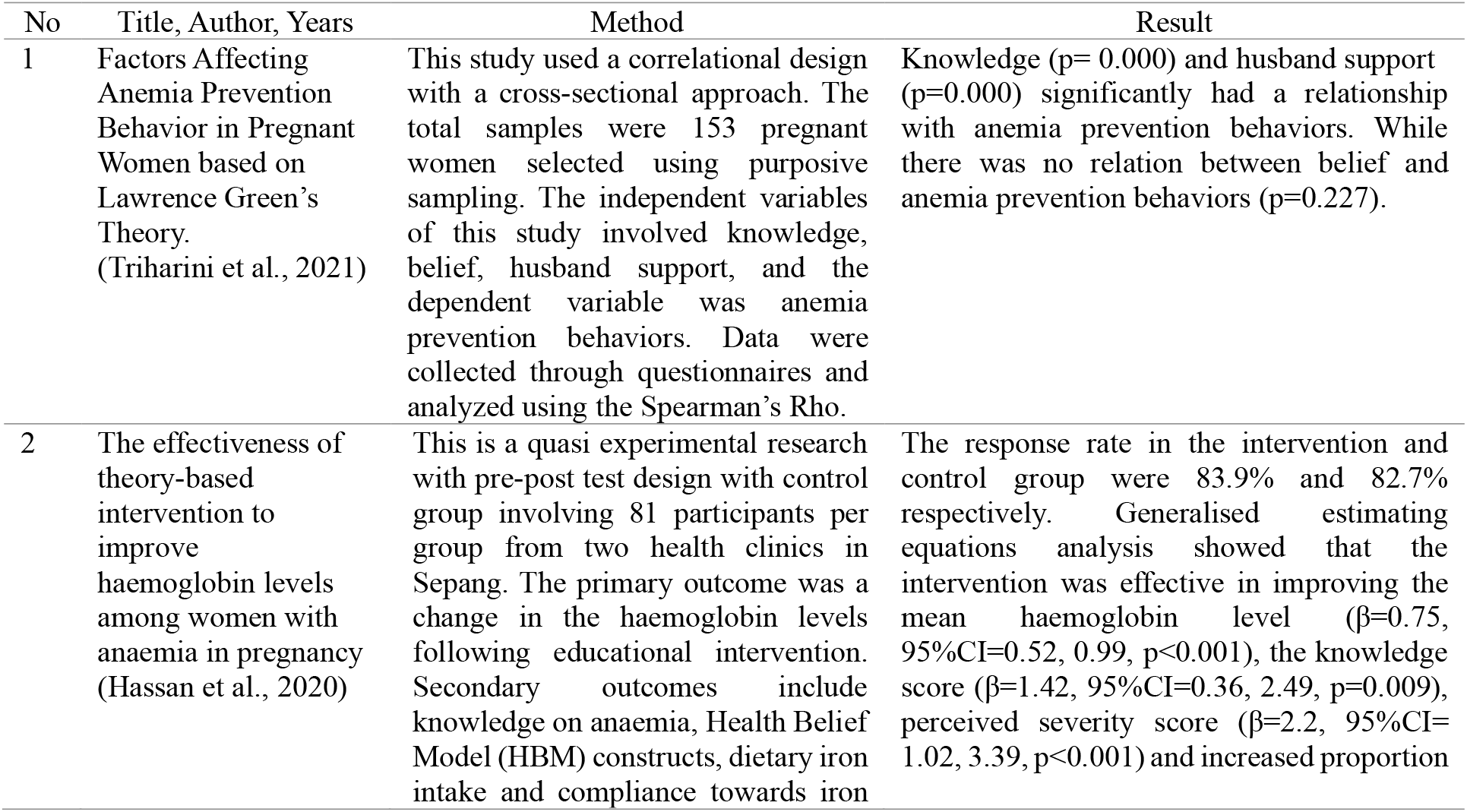
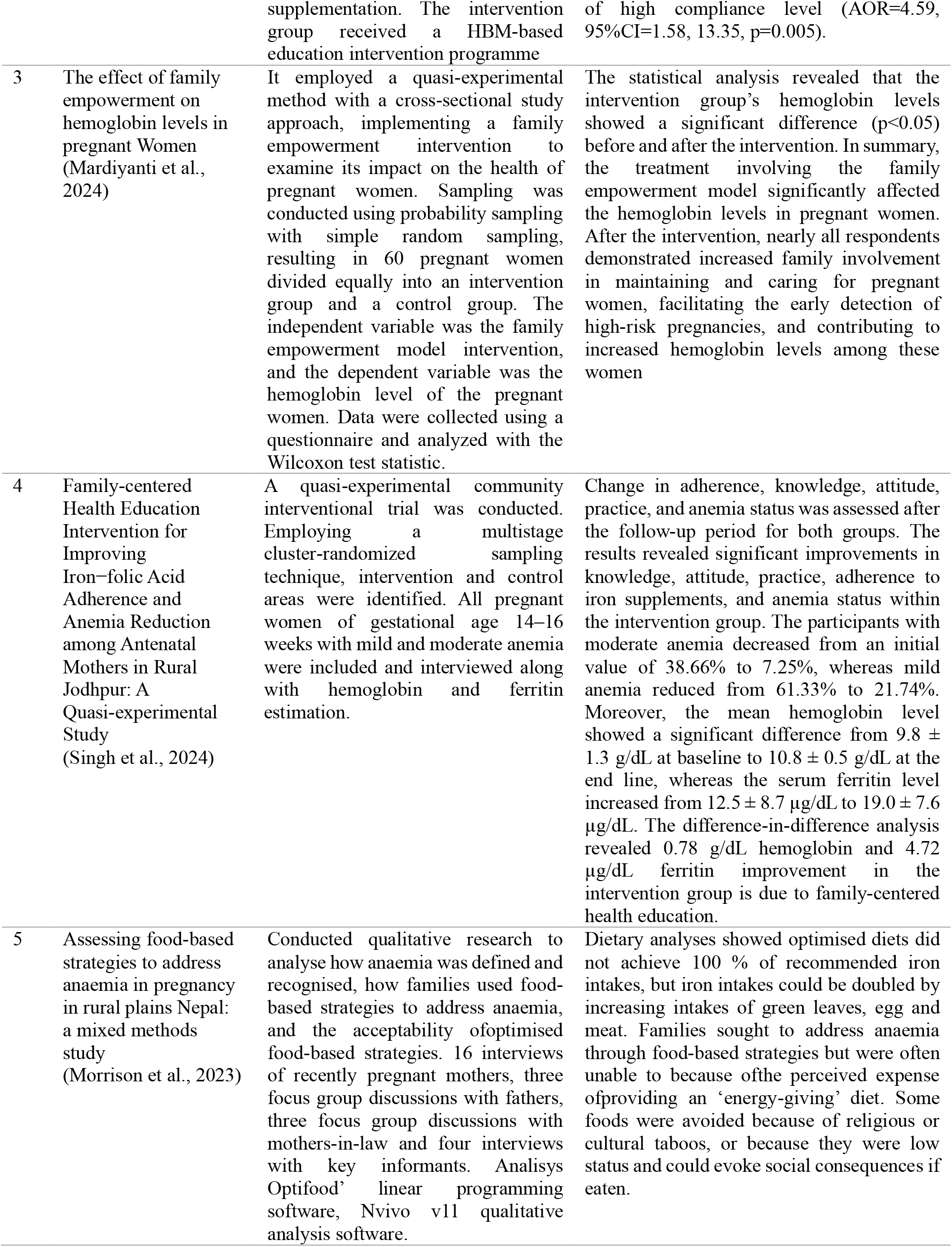
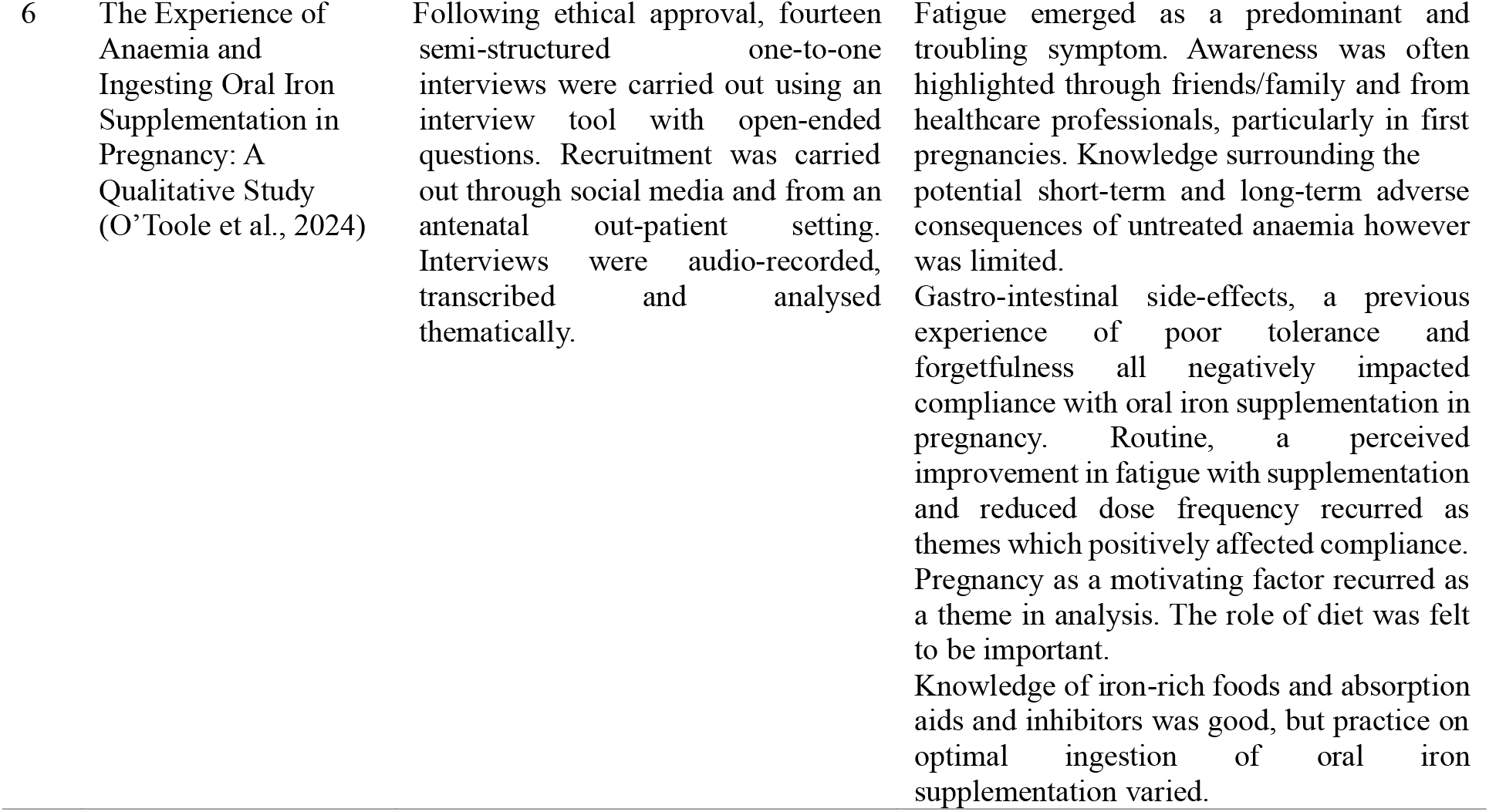
The research articles on the impact of culture-based family empowerment on anemia prevention behavior in pregnant women

## DISCUSSION

The family empowerment model has shown a significant positive impact on anemia prevention behaviors in pregnant women. This approach emphasizes the role of family support and education in improving the health outcomes. Key findings include increased hemoglobin levels,(Hassan et al., 2020; Mardiyanti et al., 2024; Singh et al., 2024; Triharini et al., 2021) increased family support,(Mardiyanti et al., 2024; Singh et al., 2024; Triharini et al., 2018) behavioral changes,(Mardiyanti et al., 2024; Morrison et al., 2023; O’Toole et al., 2024; Singh et al., 2024) and educational interventions.(Singh et al., 2024)

Table 1 shows that family empowerment interventions significantly increased the hemoglobin levels in pregnant women. For example, one study showed a significant increase in hemoglobin levels post-intervention (p<0.05).(Mardiyanti et al., 2024) Another study confirmed similar results, showing significant increases in hemoglobin levels and anemia prevention behaviors. (Singh et al., 2024)

Table 1 shows that family support plays a significant role in the effectiveness of anemia prevention strategies.(Mardiyanti et al., 2024; Singh et al., 2024; Triharini et al., 2018) Educational interventions aimed at increasing family support have been shown to significantly improve maternal anemia-preventing behaviors, such as adherence to iron supplements and intake of iron-rich foods. This is supported by the finding that a family empowerment model results in increased family involvement in caring for and nurturing pregnant women.

Table 1 shows that family support such as from a husband can encourage behavioral changes in pregnant women.^9^ Empowerment encourages behavioral changes in pregnant women by increasing their independence and commitment to preventing anemia.(Mardiyanti et al., 2024) This includes better compliance with iron supplementation.

Implementing educational intervention programs in families has been shown to be effective in changing dietary and nutritional behavior in pregnant women.(Triharini et al., 2018) These programs increase knowledge, perceptions, and practices related to anemia prevention. (Triharini et al., 2018, 2023) Integrating family empowerment with these educational interventions further increases their effectiveness.(Triharini et al., 2018, 2023)

## CONCLUSION

The family empowerment model, when combined with transcultural nursing principles, significantly impacts anemia prevention behaviors in pregnant women. This approach not only increases hemoglobin levels but also enhances family support and encourages positive behavioral changes. Health workers are encouraged to adopt these models to optimize anemia prevention strategies and improve maternal health outcomes.

## Data Availability

All data produced in the present study are available upon reasonable request to the authors
All data produced in the present work are contained in the manuscript

